# Cognitive endophenotypes in racially- and ethnically-diverse middle-aged adult offspring aggregate with parental magnetic resonance imaging biomarkers of Alzheimer’s disease risk

**DOI:** 10.1101/2021.01.15.21249832

**Authors:** Patrick J. Lao, Indira C. Turney, Justina Avila-Reiger, Jet M.J. Vonk, Miguel Arce Rentería, Dominika Seblova, Anthony G. Chesebro, Krystal K. Laing, Erica Amarante, Michelle Martinez, Judes Fleurimont, José Gutierrez, Nicole Schupf, Richard Mayeux, Jennifer J. Manly, Adam M. Brickman

## Abstract

A family history of Alzheimer’s disease (AD) increases risk for AD in an individual by 1.5-to 3-fold. Heritability of AD risk may be due in part to the aggregation of neurodegeneration and cerebrovascular changes with cognitive endophenotypes within families. The purpose of this study was to determine the extent to which cognitive functioning in middle-aged adults is associated with objectively-measured neurodegenerative and cerebrovascular neuroimaging markers linked to risk for clinical AD in their parents, and the extent to which these associations differ by race/ethnicity and language, as proxy variables for social advantage. Middle-aged children enrolled in the Offspring study (n=356; 53.1±10.1 years old; 13% Non-Hispanic White, 27% Non-Hispanic Black, 26% Latinx tested in English, 34% Latinx tested in Spanish; 65% women; 13.5±3.4 years education) were administered the NIH Toolbox, a computerized neuropsychological battery, in their preferred language. Older adults were a subset of the Washington-Heights Inwood Columbia Aging Project (77.3±6.6 years old; 75% women; 10.0±4.6 years education) who underwent T1w- and T2w-MRI and who had a child enrolled in the Offspring study. We tested the associations of parental MRI measures reflecting neurodegeneration (hippocampal volume, cortical thickness) and cerebrovascular disease (white matter hyperintensity (WMH) volume, presence of infarct) with cognitive tests scores in Offspring participants. We further stratified the models by race/ethnicity. Better offspring cognitive scores aggregated lower parental neurodegeneration and cerebrovascular disease among Non-Hispanic White and Latinx participants, and with lower parental cerebrovascular disease alone among Non-Hispanic Black participants. Associations were generally strongest in Non-Hispanic White participants compared to the other groups. These results suggest a more consistent link between offspring cognitive endophenotype and parental brain health in intergenerational AD transmission among Non-Hispanic White participants compared to racial/ethnic and minority groups in which other social factors may be adding variance.

## Introduction

The risk of late-onset Alzheimer’s disease (AD) is 1.5-3 times greater for those with a first-degree relative with AD^1, 2^, but the factors that contribute to this high degree of heritability are poorly understood. The extent to which AD risk is transmitted from one generation to the next reflects the impact of genetic and social pathways on biology and cognition across the entire lifespan^3, 4^. Intergenerational studies that examine the aggregation of neurobiological changes and cognitive phenotypes can provide insight into these pathways, particularly across racial and ethnic groups who are differentially affected by AD^5-8^. The purpose of this study was to examine whether cognitive functioning among middle-aged adults is related to neuroimaging markers of neurodegeneration and cerebrovascular disease in their parents and whether these associations differ across racial/ethnic and language groups.

Previous studies examined heritability of AD risk by determining the relationship of having a first-degree family history of AD diagnosis with various risk factors and biomarkers for AD in the proband. These efforts showed that a positive family history is related to regional brain hypometabolism on FDG-PET^9^, higher amyloid burden on PiB-PET^9^, higher amyloid and pathological tau burden in CSF^10^, higher white matter hyperintensity volume^11^, decreased temporal cortex activation on functional MRI^12^, lower brain volume on structural MRI^13^, lower white matter integrity on diffusion tensor imaging (DTI)^14^, and worse cognitive performance^15^. These studies generally relied on retrospective assessment of parental diagnostic status, which is susceptible to recall bias, and on clinic-based samples comprising primarily Non-Hispanic White participants, which do not represent the growing diversity of older adults and those disproportionately affected by the disease^5^.

In contrast, the Offspring study includes the adult, middle-aged, children of participants from the Washington Heights-Inwood Columbia Aging Project (WHICAP), a community-based, longitudinal, cohort study that is racially/ethnically and linguistically diverse, comprising Non-Hispanic White, Non-Hispanic Black/African American, and Hispanic/Latinx older adults that represent the community surrounding Columbia University Medical Center (note that we will refer to Non-Hispanic Black and Latinx throughout the text). We designed the Offspring study to examine how risk for AD is transmitted from one generation to the next through the determination of aggregation of AD endophenotypes within families, across cohorts. Taking a lifespan biospsychosocial approach^4^, the Offspring study focuses on middle aged participants for several reasons: the earliest pathological changes associated with AD can occur decades before onset of dementia^16^, midlife health and behavior are associated with late life outcomes^17, 18^, and restricting analysis to older adults only may be biased by selective survival^19^. A primary motivator of the Offspring study is to identify factors that account for the well-documented racial and ethnic disparities in AD risk^5^. We hypothesize that one source of these disparities is the more prominent role that cerebrovascular disease plays in symptom severity^20-22^ and perhaps pathogenesis^23-25^ of AD among Non-Hispanic Black and Latinx people^26^.

In the current study, we examined whether cognitive functioning in middle-aged adults is related to common magnetic resonance imaging (MRI) markers of neurodegeneration and cerebrovascular disease in their parents. We tested the hypothesis that lower cognitive functioning in midlife, which reflects an at-risk cognitive endophenotype for later AD, would be associated with neurodegenerative and cerebrovascular features in the parent generation, which can be conceptualized as biomarkers of AD risk. Furthermore, we hypothesized that the strength of these associations would vary by race/ethnicity and language group, with cerebrovascular features aggregating with cognition more strongly among Non-Hispanic Black and Latinx participants.

## Methods

### Participants

A subset of participants from the Washington Heights Inwood Columbia Aging Project (WHICAP) were investigated if (1) they participated in the MRI imaging substudy (approximately 3000 out of 8000 total WHICAP participants) and (2) they had a child enrolled in the Offspring study (approximately 2000 total Offspring participants recruited from all WHICAP families). The overlap of those two criteria was 362 parents and middle-aged adult offspring. Briefly, WHICAP is an ongoing longitudinal study that includes Medicare-eligible older adults (≥65 yrs) recruited from the community surrounding Columbia University Irving Medical Center (recruitment waves: 1992, 1999, 2009). Their adult children were contacted for participation in the Offspring study^27^. Self-reported race/ethnicity follows the 2000 Census guidelines^28^, and was performed independently for parents and offspring (i.e., there is slight mismatching across generations). We report Hispanic in WHICAP, whereas we report Latinx in Offspring due to generational differences in self-identification. We use race/ethnicity and language as a proxy for social advantage, and not as a biological or genetic variable. In WHICAP, we report sex/gender rather than sex or gender because the question was not detailed enough to determine how the participants answered; however, questionnaires were updated in Offspring to distinguish between the two, and we report sex.

### WHICAP subsample

In WHICAP, T1-weighted MRI scans were performed on a 1.5T Philips Intera scanner or a 3T Philips Achieva scanner using comparable magnetization prepared rapid acquisition gradient echo (MPRAGE) sequences (repetition time (TR)/echo time (TE): 20/2.1ms, flip angle: 20°, voxel size: 0.94×0.94×1.3mm^3^; TR/TE: 6.5/3.0ms, flip angle: 8°, voxel size: 1×1×1mm^3^, respectively), depending on their time of scan in the study. Twelve WHICAP participants were excluded for having dementia at their first imaging visit. T1-weighted MRI were anatomically segmented into regions of interest using FreeSurfer^29^ and manually edited by a team of image analysts as needed. To derive markers of neurodegeneration, we quantified hippocampal volume, adjusted for intracranial volume (ICV), and cortical thickness in an AD signature, composite region^30^, which included medial and inferior temporal, temporal pole, posterior cingulate, precuneus, and superior parietal cortices. Similarly, T2-weighted MRI scans were collected on a 1.5T Philips Intera scanner or a 3T Philips Achieva scanner using comparable fluid attenuated inversion recovery (FLAIR) sequences (TR/TE/inversion time (TI): 5500/144/1900ms, flip angle: 90°, voxel size: 0.98×0.98×3.0mm^3^; TR/TE/TI: 8000/332/2400ms, flip angle: 50°, voxel size: 1.1×1.1×0.6mm^3^, respectively). To derive markers of small vessel cerebrovascular disease, baseline T2-weighted MRI were intensity segmented into regions of hyperintensity^31^ and manually edited by a team of image analysts as needed. We quantified the WMH volume in the parietal lobe and log-transformed for normality. To identify MRI evidence of infarcts, baseline T2-weighted FLAIR MRI sequences were manually searched for discrete hypointense lesions larger than 5 mm with a partial or complete hyperintense ring and confirmed on T1-weighted images as areas of hypointensity. Because most participants had no infarcts and the range was up to ten infarcts with most having only one, we dichotomized the variable into presence of infarcts (i.e., 0 infarcts vs ≥1 infarct). Representative images of each parental imaging biomarker of AD risk is shown in Figure 1.

**Figure 1.**
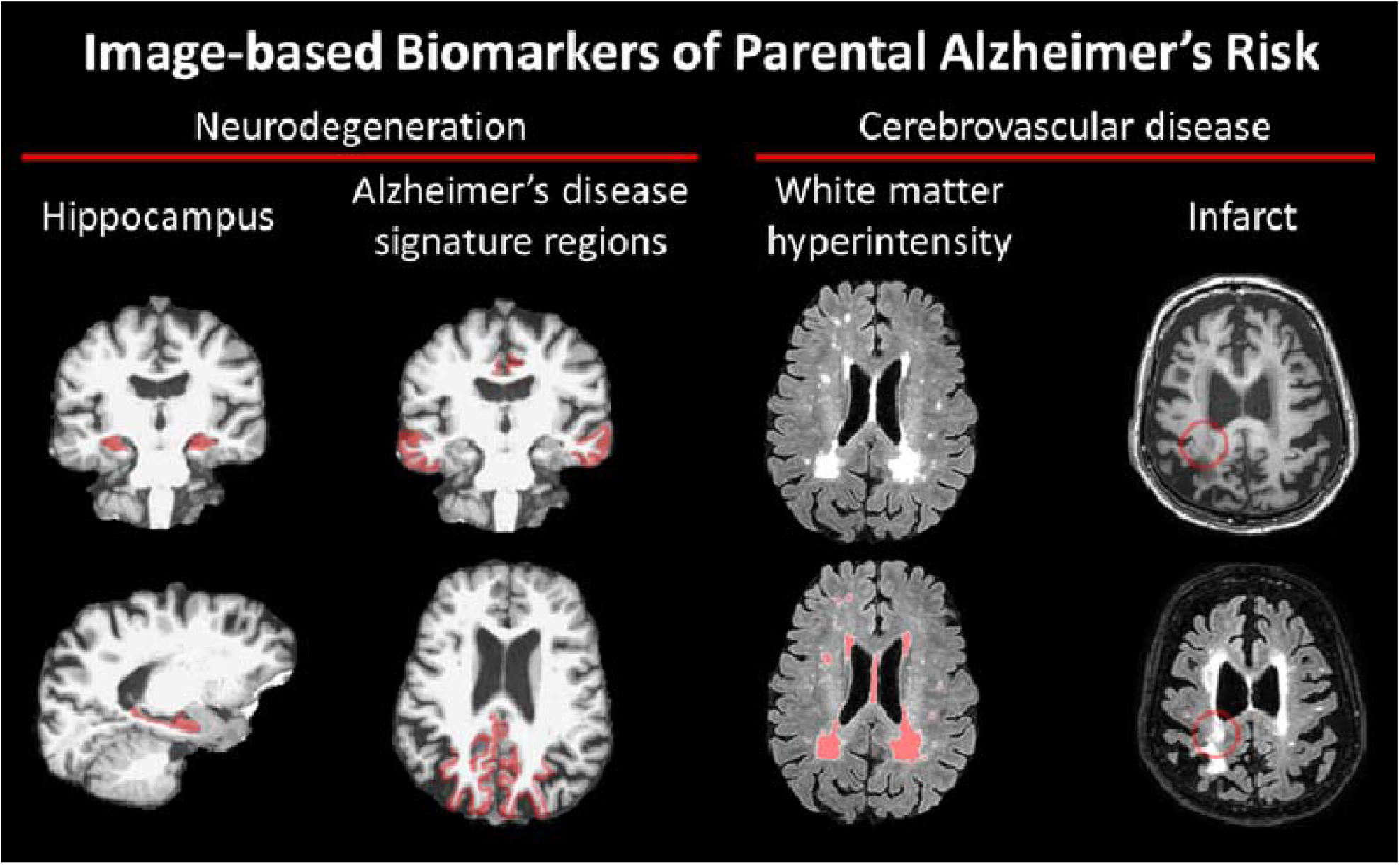
Visualization of parental imaging biomarkers of Alzheimer’s disease (AD) risk showing the hippocampus segmentation (coronal and sagittal views of T1), AD signature composite region segmentation (coronal and axial views of T1), white matter hyperintensity segmentation (axial views of T2 FLAIR), and the presence of an infarct (axial views of T1 and T2 FLAIR).

### Offspring subsample

In the Offspring study, the NIH Toolbox^32^ was administered in the participant’s language of choice (i.e., English or Spanish). NIH Toolbox is a computerized neuropsychological test battery; we included the List Sorting Test (working memory); the Dimensional Change Card Sorting Test (executive functioning), the Pattern Comparison Test (processing speed), and the Flanker Test (inhibitory control and attention). Higher raw scores on all tests reflect better performances. Multiple offspring per WHICAP parent could be included in the analysis (250 family units total; 184 parents with one offspring, 43 parents with 2 offspring, 15 parents with 3 offspring, 4 parents with 4 offspring, 2 parents with 5 offspring, 1 parent with 7 offspring, 1 parent with 8 offspring).

### Statistical models

We report demographic characteristics by study and compare by race/ethnicity using ANOVA or chi-squared tests. Prior to analyses, we fit a general linear model for the parental brain measures (hippocampal volume, as a percentage of ICV; cortical thickness in AD signature regions; log-transformed WMH volume; presence of infarcts) on parental demographic characteristics (including MRI scanner) and the offspring cognitive performance measures (raw scores of working memory, executive function, processing speed, and attention) on offspring demographic characteristics. The predicted values from these models were used in all analyses to adjust for parental and offspring demographic characteristics. First, we used separate linear mixed models (random effect = family unit) to test the relationship among each parental brain measure and each offspring cognitive performance measure (i.e., 4 cognitive measures x 4 brain measures = 16 models). We then included an interaction term with race/ethnicity and report the omnibus test statistic. Next, because statistical adjustment for race/ethnicity simply compares all groups against a referent, we tested stratified models by race/ethnicity and report associations within groups. It is important to investigate racial/ethnic and language groups (i.e., lived sociocultural experiences) to sufficiently addresses the different associations among different groups due to different contributing factors such as literacy and language use in Hispanics^33^, apolipoprotein on brain connectivity^34^, cognitive benefits of education^35^, and availability of social resources^36^. Similar interactions and stratified models were run using race/ethnicity and language groups (e.g., Latinx tested in Spanish, Latinx tested in English) and are presented in the Supplemental Materials. All statistical analyses were performed in R (v4.0.2^37^).

## Results

Demographic characteristics of the included WHICAP and Offspring study participants are provided in Table 1. Most participants were Latinx, followed by Non-Hispanic Black and Non-Hispanic White, respectively. WHICAP included a higher percentage of women compared to Offspring, and parents had less education compared to middle-aged adult children. Within the WHICAP subsample, there were demographic differences among race/ethnicity groups in age (Hispanic > Non-Hispanic Black), sex/gender distribution (Non-Hispanic Black > Non-Hispanic White, Hispanic), and years of formal education (Non-Hispanic White > Non-Hispanic Black > Hispanic). Within the Offspring study subsample, there were demographic differences among race/ethnicity in years of education (Non-Hispanic White > Non-Hispanic Black > Latinx), but not in age or sex distribution.

**Table 1.**
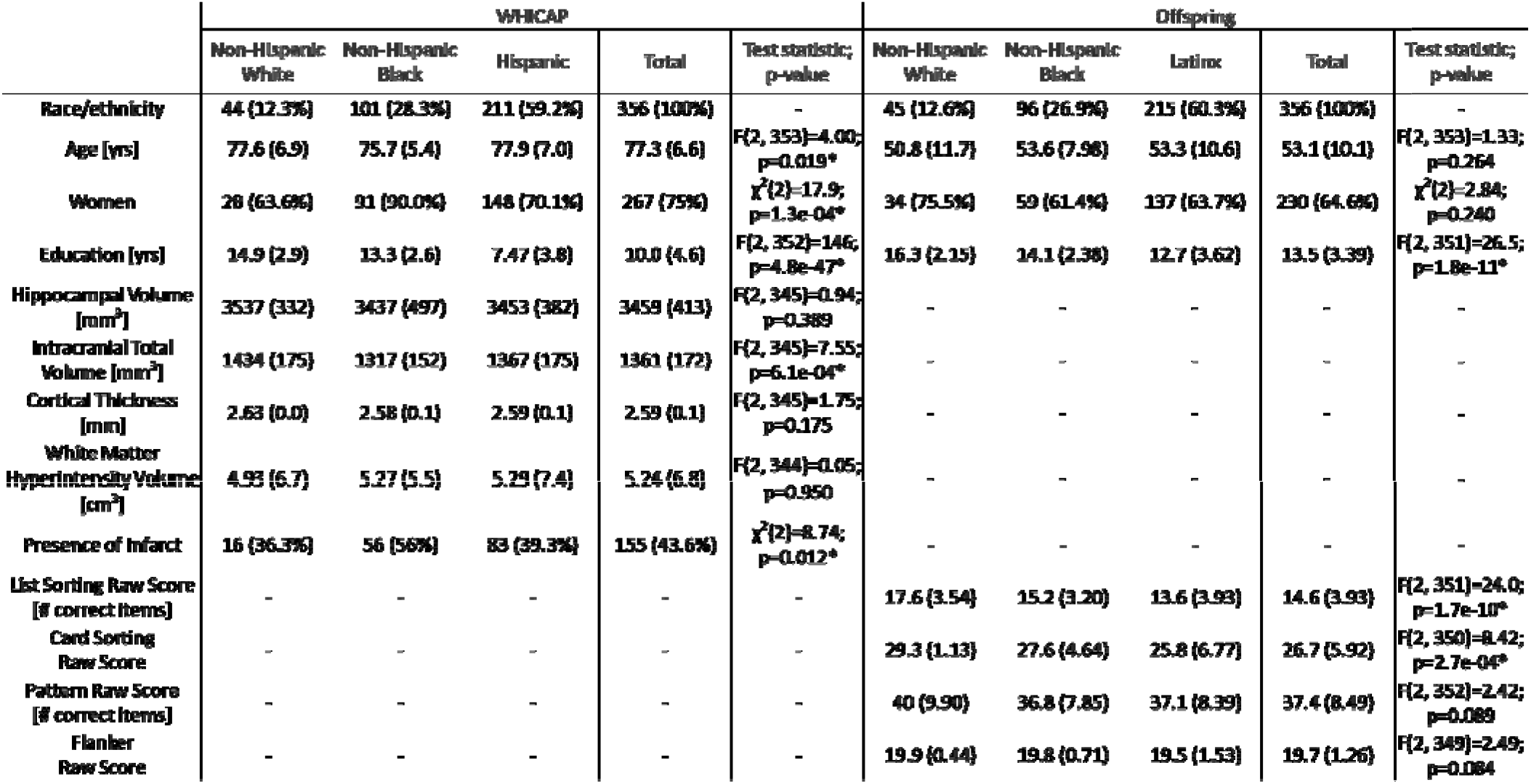
Demographic characteristics of parents with imaging and offspring with NIH Toolbox variables. Count variables are reported as number (percentage), while continuous variables are reported as mean (standard deviation). Test statistics across race/ethnicity and language groups and associated p-values are reported. An ∗ indicates p<0.05.

|  | WHICAP |  |  |  |  | Offspring |  |  |  |  |
| --- | --- | --- | --- | --- | --- | --- | --- | --- | --- | --- |
|  | Non-Hispanic White | Non-Hispanic Black | Hispanic | Total | Test statistic; p-value | Non-Hispanic White | Non-Hispanic Black | Latino | Total | Test statistic; p-value |
| Race/ethnicity | 44 (12.3%) | 101 (28.3%) | 211 (59.2%) | 356 (100%) | - | 45 (12.6%) | 96 (26.9%) | 215 (60.3%) | 356 (100%) | - |
| Age [yrs] | 77.6 (6.9) | 75.7 (5.4) | 77.9 (7.0) | 77.3 (6.6) | $F(2, 353)=4.00$; $p=0.019^*$ | 50.8 (11.7) | 53.6 (7.98) | 53.3 (10.6) | 53.1 (10.1) | $F(2, 353)=1.33$; $p=0.264$ |
| Women | 28 (63.6%) | 91 (90.0%) | 148 (70.1%) | 267 (75%) | $\chi^2(2)=17.9$; $p=1.3e-04^*$ | 34 (75.5%) | 59 (61.4%) | 137 (63.7%) | 230 (64.6%) | $\chi^2(2)=2.84$; $p=0.240$ |
| Education [yrs] | 14.9 (2.9) | 13.3 (2.6) | 7.47 (3.8) | 10.0 (4.6) | $F(2, 352)=146$; $p=4.8e-47^*$ | 16.3 (2.15) | 14.1 (2.38) | 12.7 (3.62) | 13.5 (3.39) | $F(2, 351)=26.5$; $p=1.8e-11^*$ |
| Hippocampal Volume [mm<sup>3</sup>] | 3537 (332) | 3437 (497) | 3453 (382) | 3459 (413) | $F(2, 345)=0.94$; $p=0.389$ | - | - | - | - | - |
| Intracranial Total Volume [mm<sup>3</sup>] | 1434 (175) | 1317 (152) | 1367 (175) | 1361 (172) | $F(2, 345)=7.55$; $p=6.1e-04^*$ | - | - | - | - | - |
| Cortical Thickness [mm] | 2.63 (0.0) | 2.58 (0.1) | 2.59 (0.1) | 2.59 (0.1) | $F(2, 345)=1.75$; $p=0.175$ | - | - | - | - | - |
| White Matter Hyperintensity Volume [cm<sup>3</sup>] | 4.93 (6.7) | 5.27 (5.5) | 5.29 (7.4) | 5.24 (6.8) | $F(2, 344)=0.05$; $p=0.950$ | - | - | - | - | - |
| Presence of Infarct | 16 (36.3%) | 56 (56%) | 83 (39.3%) | 155 (43.6%) | $\chi^2(2)=8.74$; $p=0.012^*$ | - | - | - | - | - |
| List Sorting Raw Score [# correct items] | - | - | - | - | - | 17.6 (3.54) | 15.2 (3.20) | 13.6 (3.93) | 14.6 (3.93) | $F(2, 351)=24.0$; $p=1.7e-10^*$ |
| Card Sorting Raw Score | - | - | - | - | - | 29.3 (1.13) | 27.6 (4.64) | 25.8 (6.77) | 26.7 (5.92) | $F(2, 350)=8.42$; $p=2.7e-04^*$ |
| Pattern Raw Score [# correct items] | - | - | - | - | - | 40 (9.90) | 36.8 (7.85) | 37.1 (8.39) | 37.4 (8.49) | $F(2, 352)=2.42$; $p=0.089$ |
| Flanker Raw Score | - | - | - | - | - | 19.9 (0.44) | 19.8 (0.71) | 19.5 (1.53) | 19.7 (1.26) | $F(2, 349)=2.49$; $p=0.084$ |

The results of familial aggregation of cognitive endophenotypes and biomarkers of neurodegeneration and cerebrovascular disease are presented in Table 2. Across all participants in this subset of the WHICAP and Offspring studies, middle-aged adult offspring had better working memory, executive function, processing speed, and attention scores if they had a parent with higher hippocampal volume and higher cortical thickness. Middle-aged offspring also had better processing speed and attention scores if their parent had lower WMH volume and no infarcts. In stratified models (Table 2), associations among offspring cognition across tests and parental neurodegeneration and cerebrovascular biomarkers were observed in Non-Hispanic White and Latinx participants, while associations among offspring cognition across tests and parental cerebrovascular biomarkers were observed in Non-Hispanic Black participants. The associations between offspring cognition in all tests and parental biomarkers were strongest in Non-Hispanic White participants compared to the other groups, with the exception that associations between offspring cognition in all domains and parental cortical thickness were similarly stronger in Non-Hispanic White and Latinx participants compared to Non-Hispanic Black participants (Figure 2, Table 3).

**Table 2.**
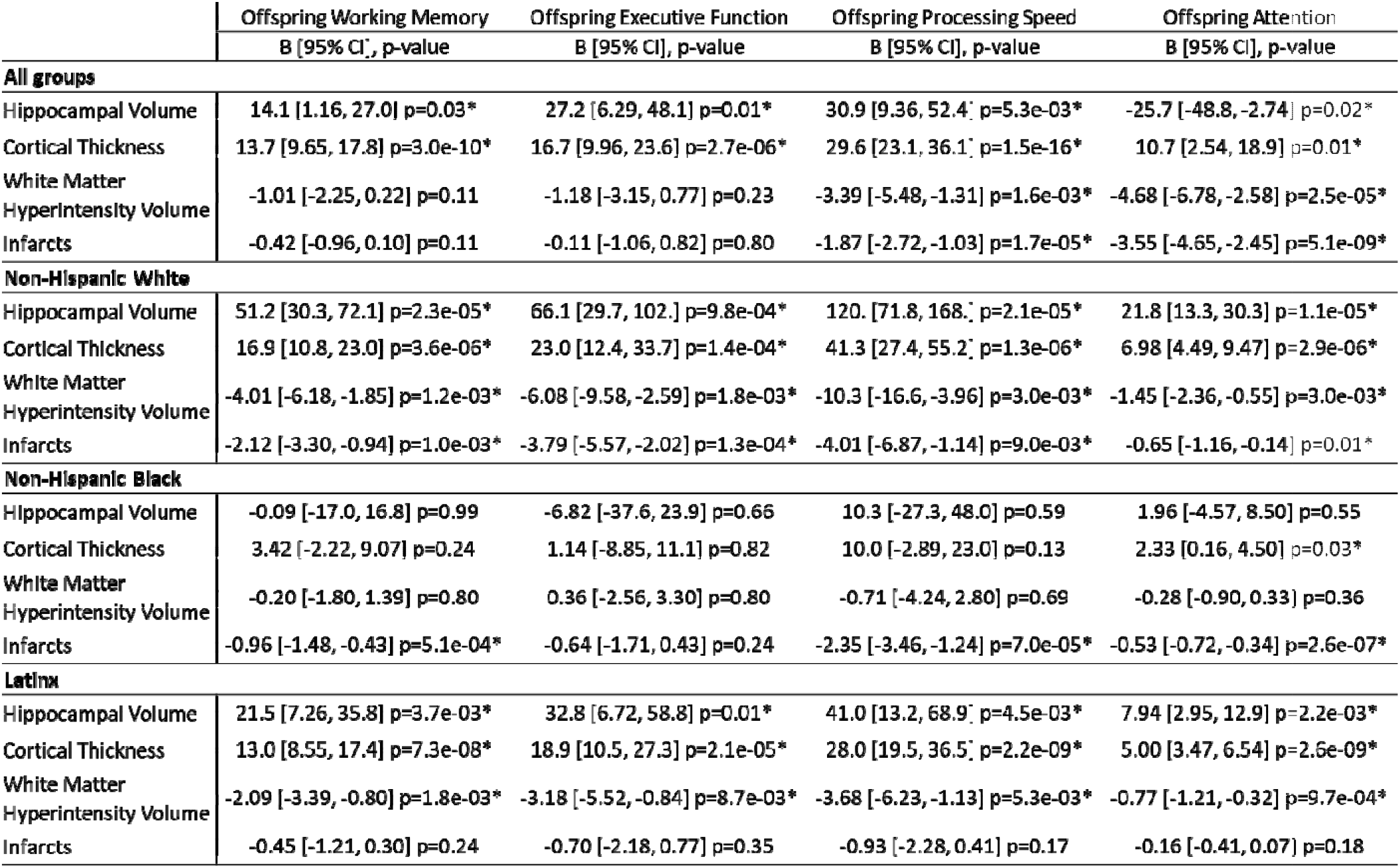
Associations of parental brain biomarkers and offspring cognitive performance. Separate models were run for each parental brain biomarker and each offspring cognitive test score in all groups and the same models stratified in each race/ethnicity group. An ∗ represents p<0.05 in the overall models or the stratified models.

|  | Offspring Working Memory<br>B [95% CI], p-value | Offspring Executive Function<br>B [95% CI], p-value | Offspring Processing Speed<br>B [95% CI], p-value | Offspring Attention<br>B [95% CI], p-value |
| --- | --- | --- | --- | --- |
| **All groups** |  |  |  |  |
| Hippocampal Volume | 14.1 [1.16, 27.0] $p=0.03^*$ | 27.2 [6.29, 48.1] $p=0.01^*$ | 30.9 [9.36, 52.4] $p=5.3e-03^*$ | -25.7 [-48.8, -2.74] $p=0.02^*$ |
| Cortical Thickness | 13.7 [9.65, 17.8] $p=3.0e-10^*$ | 16.7 [9.96, 23.6] $p=2.7e-06^*$ | 29.6 [23.1, 36.1] $p=1.5e-16^*$ | 10.7 [2.54, 18.9] $p=0.01^*$ |
| White Matter Hyperintensity Volume | -1.01 [-2.25, 0.22] $p=0.11$ | -1.18 [-3.15, 0.77] $p=0.23$ | -3.39 [-5.48, -1.31] $p=1.6e-03^*$ | -4.68 [-6.78, -2.58] $p=2.5e-05^*$ |
| Infarcts | -0.42 [-0.96, 0.10] $p=0.11$ | -0.11 [-1.06, 0.82] $p=0.80$ | -1.87 [-2.72, -1.03] $p=1.7e-05^*$ | -3.55 [-4.65, -2.45] $p=5.1e-09^*$ |
| **Non-Hispanic White** |  |  |  |  |
| Hippocampal Volume | 51.2 [30.3, 72.1] $p=2.3e-05^*$ | 66.1 [29.7, 102.] $p=9.8e-04^*$ | 120. [71.8, 168.] $p=2.1e-05^*$ | 21.8 [13.3, 30.3] $p=1.1e-05^*$ |
| Cortical Thickness | 16.9 [10.8, 23.0] $p=3.6e-06^*$ | 23.0 [12.4, 33.7] $p=1.4e-04^*$ | 41.3 [27.4, 55.2] $p=1.3e-06^*$ | 6.98 [4.49, 9.47] $p=2.9e-06^*$ |
| White Matter Hyperintensity Volume | -4.01 [-6.18, -1.85] $p=1.2e-03^*$ | -6.08 [-9.58, -2.59] $p=1.8e-03^*$ | -10.3 [-16.6, -3.96] $p=3.0e-03^*$ | -1.45 [-2.36, -0.55] $p=3.0e-03^*$ |
| Infarcts | -2.12 [-3.30, -0.94] $p=1.0e-03^*$ | -3.79 [-5.57, -2.02] $p=1.3e-04^*$ | -4.01 [-6.87, -1.14] $p=9.0e-03^*$ | -0.65 [-1.16, -0.14] $p=0.01^*$ |
| **Non-Hispanic Black** |  |  |  |  |
| Hippocampal Volume | -0.09 [-17.0, 16.8] $p=0.99$ | -6.82 [-37.6, 23.9] $p=0.66$ | 10.3 [-27.3, 48.0] $p=0.59$ | 1.96 [-4.57, 8.50] $p=0.55$ |
| Cortical Thickness | 3.42 [-2.22, 9.07] $p=0.24$ | 1.14 [-8.85, 11.1] $p=0.82$ | 10.0 [-2.89, 23.0] $p=0.13$ | 2.33 [0.16, 4.50] $p=0.03^*$ |
| White Matter Hyperintensity Volume | -0.20 [-1.80, 1.39] $p=0.80$ | 0.36 [-2.56, 3.30] $p=0.80$ | -0.71 [-4.24, 2.80] $p=0.69$ | -0.28 [-0.90, 0.33] $p=0.36$ |
| Infarcts | -0.96 [-1.48, -0.43] $p=5.1e-04^*$ | -0.64 [-1.71, 0.43] $p=0.24$ | -2.35 [-3.46, -1.24] $p=7.0e-05^*$ | -0.53 [-0.72, -0.34] $p=2.6e-07^*$ |
| **Latinx** |  |  |  |  |
| Hippocampal Volume | 21.5 [7.26, 35.8] $p=3.7e-03^*$ | 32.8 [6.72, 58.8] $p=0.01^*$ | 41.0 [13.2, 68.9] $p=4.5e-03^*$ | 7.94 [2.95, 12.9] $p=2.2e-03^*$ |
| Cortical Thickness | 13.0 [8.55, 17.4] $p=7.3e-08^*$ | 18.9 [10.5, 27.3] $p=2.1e-05^*$ | 28.0 [19.5, 36.5] $p=2.2e-09^*$ | 5.00 [3.47, 6.54] $p=2.6e-09^*$ |
| White Matter Hyperintensity Volume | -2.09 [-3.39, -0.80] $p=1.8e-03^*$ | -3.18 [-5.52, -0.84] $p=8.7e-03^*$ | -3.68 [-6.23, -1.13] $p=5.3e-03^*$ | -0.77 [-1.21, -0.32] $p=9.7e-04^*$ |
| Infarcts | -0.45 [-1.21, 0.30] $p=0.24$ | -0.70 [-2.18, 0.77] $p=0.35$ | -0.93 [-2.28, 0.41] $p=0.17$ | -0.16 [-0.41, 0.07] $p=0.18$ |

**Table 3.**
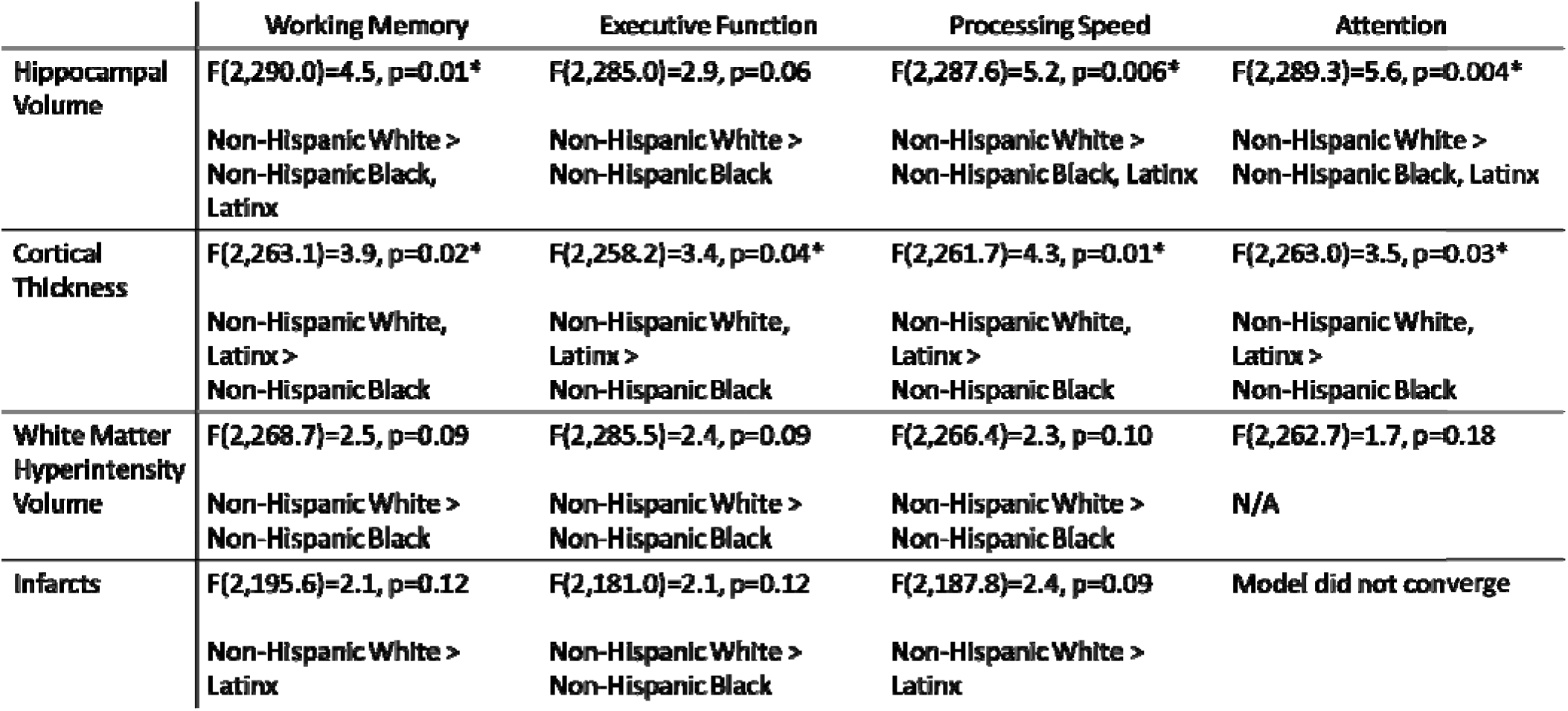
Interactions by race/ethnicity on parental brain measures with offspring cognition. As an example of the notation, X > W, Y, Z indicates that group X had a stronger association (in either the positive or negative direction; see Table 2 for association direction and magnitude) compared to group W, group Y, and group Z, while groups W, Y, and Z did not differ from each other. If a group is not listed, they were most likely intermediate between groups and were not different from any other groups.

|  | **Working Memory** | **Executive Function** | **Processing Speed** | **Attention** |
| --- | --- | --- | --- | --- |
| **Hippocampal Volume** | **F(2,290.0)=4.5, p=0.01\***<br>**Non-Hispanic White > Non-Hispanic Black, Latinx** | **F(2,285.0)=2.9, p=0.06**<br>**Non-Hispanic White > Non-Hispanic Black** | **F(2,287.6)=5.2, p=0.006\***<br>**Non-Hispanic White > Non-Hispanic Black, Latinx** | **F(2,289.3)=5.6, p=0.004\***<br>**Non-Hispanic White > Non-Hispanic Black, Latinx** |
| **Cortical Thickness** | **F(2,263.1)=3.9, p=0.02\***<br>**Non-Hispanic White, Latinx > Non-Hispanic Black** | **F(2,258.2)=3.4, p=0.04\***<br>**Non-Hispanic White, Latinx > Non-Hispanic Black** | **F(2,261.7)=4.3, p=0.01\***<br>**Non-Hispanic White, Latinx > Non-Hispanic Black** | **F(2,263.0)=3.5, p=0.03\***<br>**Non-Hispanic White, Latinx > Non-Hispanic Black** |
| **White Matter Hyperintensity Volume** | **F(2,268.7)=2.5, p=0.09**<br>**Non-Hispanic White > Non-Hispanic Black** | **F(2,285.5)=2.4, p=0.09**<br>**Non-Hispanic White > Non-Hispanic Black** | **F(2,266.4)=2.3, p=0.10**<br>**Non-Hispanic White > Non-Hispanic Black** | **F(2,262.7)=1.7, p=0.18**<br>**N/A** |
| **Infarcts** | **F(2,195.6)=2.1, p=0.12**<br>**Non-Hispanic White > Latinx** | **F(2,181.0)=2.1, p=0.12**<br>**Non-Hispanic White > Non-Hispanic Black** | **F(2,187.8)=2.4, p=0.09**<br>**Non-Hispanic White > Latinx** | **Model did not converge** |

**Figure 2.**
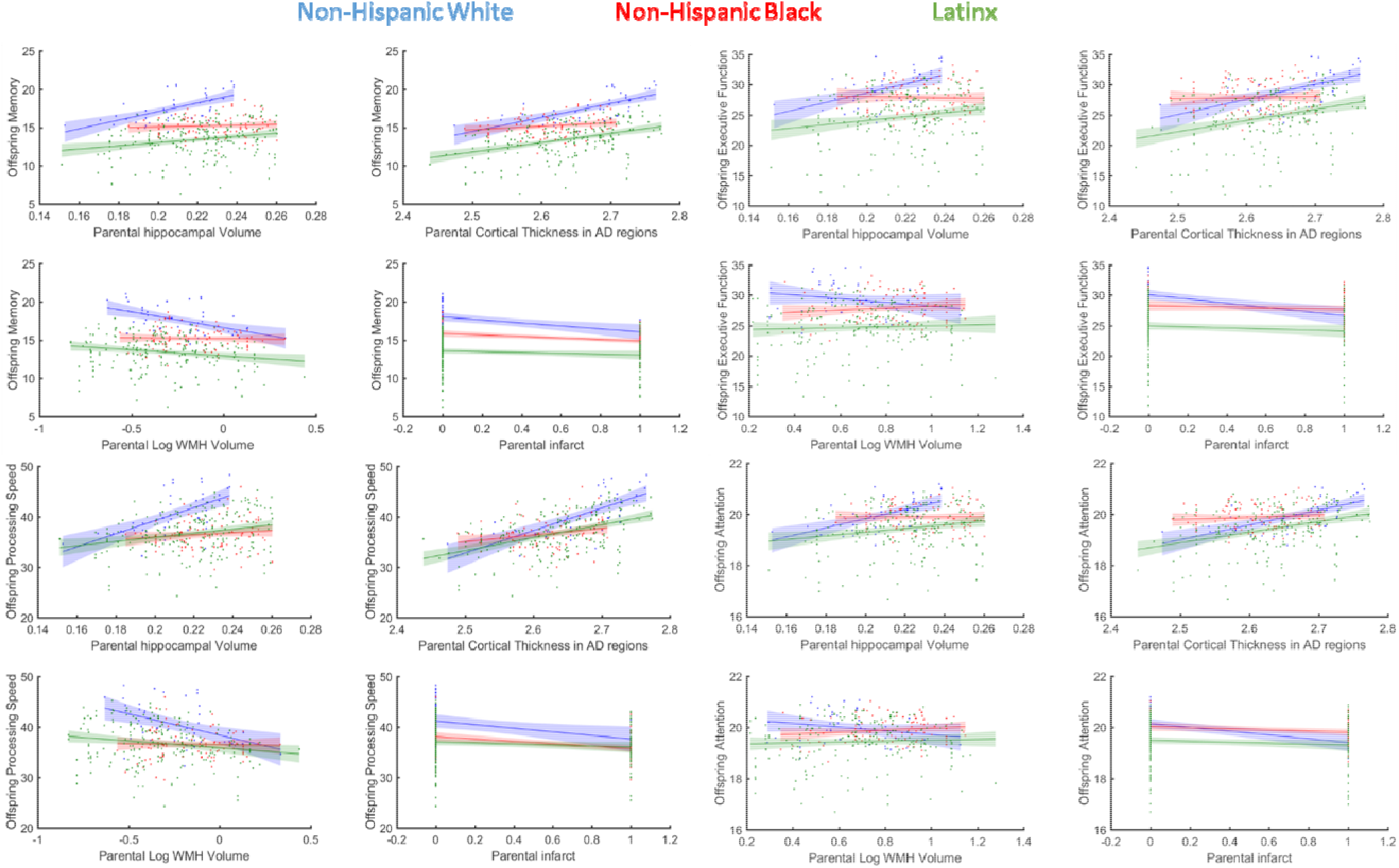
The association of offspring attention scores with parental brain measures by race/ethnicity.

Further considering preferred testing language in Latinx offspring, there were demographic differences among race/ethnicity and language groups in age (Latinx tested in Spanish > Non-Hispanic Black > Latinx tested in English; Latinx tested in Spanish > Non-Hispanic White) and years of education (Non-Hispanic White > Non-Hispanic Black, Latinx tested in English > Latinx tested in Spanish), but not in sex distribution (Supplemental Table 1). In language stratified models (Supplemental Table 2; Supplemental Figure 1), associations among offspring cognition across tests and parental neurodegeneration biomarkers were observed in Latinx participants tested in English and Spanish, while associations among offspring cognition across tests and parental cerebrovascular disease biomarkers were most apparent in Latinx participants tested in English. All significant associations were found to be in the hypothesized direction (i.e., better offspring cognition aggregated with lower parental neurodegeneration and cerebrovascular disease) except for the relationships between offspring executive function and attention scores and parental infarcts among Latinx participants tested in English.

## Conclusion

Currently, the mechanisms and pathways by which AD risk is transferred from one generation to the next is unknown, but there are likely many factors (e.g., genetic, social, environmental) that contribute to intergenerational AD risk transmission^38^. Our work characterized the familial aggregation of neurodegenerative and cerebrovascular biomarkers in parents and cognitive endophenotypes in adult children in a diverse, community-based sample to better understand the pathways for intergenerational AD risk transmission. Health disparities are due to injustice such that groups that persistently experience social disadvantage or discrimination systematically have worse health^39, 40^; therefore we investigated differences by race/ethnicity. Health disparities can exist as a difference in explanatory variable (i.e., neurodegeneration, cerebrovascular disease), as a difference in outcome (i.e., cognition), or as a difference in the mechanistic link between the two^41^. Parental hippocampal volume and cortical thickness in AD signature regions were greatest in Non-Hispanic White participants, while parental WMH volume was greater and the presence of parental infarcts was more likely in Non-Hispanic Black and Latinx participants. While some differences did not reach statistical significance in this subsample, WHICAP studies that included more participants with available neuroimaging data showed the robustness of these disparities^42, 43^. Additionally, Non-Hispanic White offspring participants generally had the highest cognitive test scores, perhaps due to their higher number of years of formal education. We also found evidence for racial/ethnic and language group differences in the association strength between exposures and outcomes, such that there was a greater association between all cognitive scores and both neurodegenerative and cerebrovascular biomarkers in Non-Hispanic White participants compared to racial/ethnic and language minority groups, suggesting that other factors beyond brain health may be adding variance in minority groups. Neurodegeneration is non-specific, and even in AD-associated regions, could be due to other lifecourse social factors, such as chronic stress due to racism^44^ or uncontrolled vascular risk factors due to lower access to healthcare^45^, and therefore may not be an appropriate biomarker of AD risk in certain groups.

Our current work extends known intra-individual associations among cognition and biomarkers of neurodegeneration and cerebrovascular disease to the intergenerational transmission of AD risk. In previous work, hippocampal volume was related to memory performance in Non-Hispanic White older adults, but not in Non-Hispanic Black or Hispanic older adults^26^. Previous studies have also found more aggressive cognitive decline^46, 47^ and a higher likelihood of developing AD^48, 49^ in those with greater WMH volume, and faster cognitive decline with greater WMH, above and beyond amyloid and neurodegeneration, in Non-Hispanic Black older adults, but not Non-Hispanic White or Hispanic older adults^47^. There is evidence that parietal WMH play a more mechanistic role in AD development and/or progression from late-onset AD^23^, autosomal dominant AD^24^ and AD in adults with Down syndrome^25^, possibly owing to the fact that the posterior cerebral artery perfuses both the parietal and temporal cortices and small vessel disease in the parietal lobe may indicate downstream damage in temporal lobe structures such as the entorhinal cortex^50^. Offspring cognitive endophenotypes aggregated with parental imaging biomarkers of AD risk most strongly in Non-Hispanic White participants, likely reflecting the homogeneity of this group compared to other groups. There were some unexpected findings, such as the aggregation of parental infarcts and higher executive function and attention scores in Latinx participants tested in English, which require further investigation and may be best understood with longitudinal data.

Limitations include the cross-sectional design and the relatively small sample size of the imaging subset in WHICAP with middle-aged children that have participated in the first wave of the Offspring study. The cross-sectional nature of these associations simply demonstrates the aggregation of parental neurodegeneration and cerebrovascular biomarkers and offspring cognitive phenotypes, but does not allow us to conclude that parental brain health causes offspring cognitive performance. It is important to consider how secular trends in improved educational opportunities and attainment, advancements in healthcare, and increases in general health awareness and health behaviors between generations might confound the aggregation of parental neurodegeneration and cerebrovascular biomarkers and offspring cognitive endophenotypes, particularly in the Latinx participants who had more schooling than their parents and were more comfortable being tested in English. Sample size might have underpowered our analyses, and results require further investigation and validation, particularly when considering all parental brain biomarkers together or other cerebrovascular measures (e.g., microbleeds, enlarged perivascular spaces). The strengths of this study include the racial and ethnic diversity and objective measures of AD risk in intergenerational cohorts.

In conclusion, our study has provided the first insights into the familial aggregation of worse levels of offspring cognition (i.e., a cognitive endophenotype of AD risk) and worse levels of parental neurodegeneration and cerebrovascular disease (i.e., brain biomarkers of AD risk), most strongly in Non-Hispanic White participants but still to a lesser degree in Non-Hispanic Black participants and Latinx participants. Future work in these ongoing imaging substudies needs to identify mediating factors for these observed familial aggregations of brain and cognitive health, potentially including cognitive test performance beyond the NIH toolbox (i.e., memory-specific tasks) and traditional AD biomarkers (i.e., amyloid-β plaques, pathological tau). Importantly, while no AD treatments exist, cerebrovascular disease can be improved through medication, and the public health impact of vascular risk factors can be lessened by socially equitable economic and environmental policies, reducing preventable pathways for intergenerational transmission of AD risk, particularly in at-risk participants.

## Data Availability

Data from the WHICAP or Offspring studies can be requested.

https://www.cumc.columbia.edu/adrc/investigators

## Acknowledgements

Data collection and sharing for this project was supported by the Washington Heights-Inwood Columbia Aging Project (WHICAP, P01AG07232, R01AG037212, RF1AG054023, R56AG034189, R01AG034189, R01AG054520) and the Offspring study (RF1 AG054070, R01 AG058067) funded by the National Institute on Aging (NIA). This manuscript was also supported by K99AG065506 and has been reviewed by WHICAP and Offspring investigators for scientific content and consistency of data interpretation with previous WHICAP and Offspring Study publications. We acknowledge the WHICAP and Offspring study participants and the WHICAP and Offspring research and support staff for their contributions to this study.

---

**Supplemental Table 1.** Demographic characteristics of parents with imaging and offspring with NIH Toolbox variables. Count variables are reported as number (percentage), while continuous variables are reported as mean (standard deviation). Test statistics across race/ethnicity and language groups and associated p-values are reported. An ∗ indicates p<0.05.

|  | WHICAP |  |  |  |  | Offspring |  |  |  |  | Test statistic; p-value |
| --- | --- | --- | --- | --- | --- | --- | --- | --- | --- | --- | --- |
|  | Non-Hispanic White | Non-Hispanic Black | Hispanic | Total | Test statistic; p-value | Non-Hispanic White | Non-Hispanic Black | Latina [English] | Latina [Spanish] | Total |  |
| Race/ethnicity and language group | 44 (12.3%) | 101 (28.3%) | 211 (59.2%) | 356 (100%) | - | 45 (12.6%) | 96 (26.9%) | 93 (26.1%) | 122 (34.2%) | 356 (100%) | - |
| Age [yrs] | 77.6 (6.9) | 75.7 (5.4) | 77.9 (7.0) | 77.3 (6.6) | $F(2, 353)=4.00$; $p=0.019^*$ | 50.8 (11.7) | 53.6 (7.98) | 47.7 (9.21) | 57.6 (9.62) | 53.1 (10.1) | $F(3, 352)=20.7$; $p=2.2e-12^*$ |
| Women | 28 (63.6%) | 91 (90.0%) | 148 (70.1%) | 267 (75%) | $\chi^2(2)=17.9$; $p=1.3e-04^*$ | 34 (75.5%) | 59 (61.4%) | 53 (56.9%) | 84 (68.8%) | 230 (64.6%) | $\chi^2(3)=6.09$; $p=0.106$ |
| Education [yrs] | 14.9 (2.9) | 13.3 (2.6) | 7.47 (3.8) | 10.0 (4.6) | $F(2, 352)=146$; $p=4.8e-47^*$ | 16.3 (2.15) | 14.1 (2.38) | 14.3 (2.45) | 11.4 (3.87) | 13.5 (3.39) | $F(3, 350)=37.0$; $p=7.7e-21^*$ |
| Hippocampal Volume [mm<sup>3</sup>] | 3537 (332) | 3437 (497) | 3453 (382) | 3459 (413) | $F(2, 345)=0.94$; $p=0.389$ | - | - | - | - | - | - |
| Intracranial Total Volume [mm<sup>3</sup>] | 1434 (175) | 1317 (152) | 1367 (175) | 1361 (172) | $F(2, 345)=7.55$; $p=6.1e-04^*$ | - | - | - | - | - | - |
| Cortical Thickness [mm] | 2.63 (0.0) | 2.58 (0.1) | 2.59 (0.1) | 2.59 (0.1) | $F(2, 345)=1.75$; $p=0.175$ | - | - | - | - | - | - |
| White Matter Hyperintensity Volume [cm<sup>3</sup>] | 4.93 (6.7) | 5.27 (5.5) | 5.29 (7.4) | 5.24 (6.8) | $F(2, 344)=0.05$; $p=0.950$ | - | - | - | - | - | - |
| Presence of Infarct | 16 (36.3%) | 56 (56%) | 83 (39.3%) | 155 (43.6%) | $\chi^2(2)=8.74$; $p=0.012^*$ | - | - | - | - | - | - |
| List Sorting Raw Score [/# correct items] | - | - | - | - | - | 17.6 (3.54) | 15.2 (3.20) | 15.7 (3.30) | 12.0 (3.63) | 14.6 (3.93) | $F(3, 350)=39.0$; $p=8.7e-22^*$ |
| Card Sorting Raw Score | - | - | - | - | - | 29.3 (1.13) | 27.6 (4.64) | 28.2 (3.56) | 23.8 (7.98) | 26.7 (5.92) | $F(3, 349)=17.0$; $p=2.5e-10^*$ |
| Pattern Raw Score [/# correct items] | - | - | - | - | - | 40 (9.30) | 36.8 (7.85) | 39.4 (8.47) | 35.3 (7.91) | 37.4 (8.40) | $F(3, 351)=5.82$; $p=6.8e-04^*$ |
| Flanker Raw Score | - | - | - | - | - | 19.9 (0.44) | 19.8 (0.71) | 19.9 (0.31) | 19.2 (1.99) | 19.7 (1.26) | $F(3, 348)=7.11$; $p=1.2e-04^*$ |

**Supplemental Table 2.**
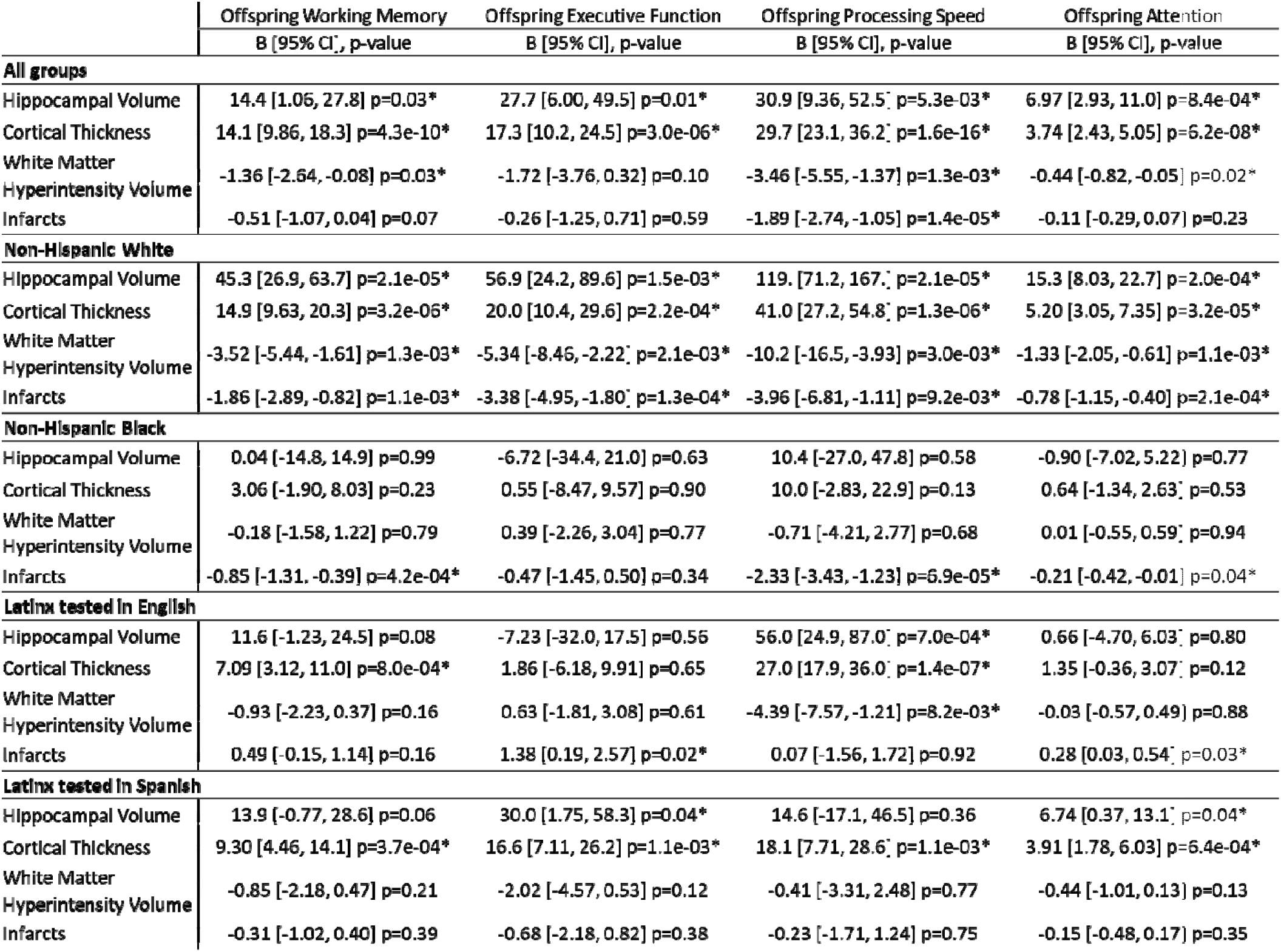
Associations of parental brain biomarkers and offspring cognitive performance. Separate models were run for each parental brain biomarker and each offspring cognitive test score in all groups and the same models stratified in each race/ethnicity language group. An ∗ represents p<0.05 in the overall models or the stratified models.

**Supplemental Table 3.** Interactions by race/ethnicity and language groups on parental brain measures with offspring cognition. As an example of the notation, X > W, Y, Z indicates that group X had a stronger association (in either the positive or negative direction; see Supplemental Table 2 for association direction and magnitude) compared to group W, group Y, and group Z, while groups W, Y, and Z did not differ from each other. If a group is not listed, they were most likely intermediate between groups and were not different from any other groups.

|  | Working Memory | Executive Function | Processing Speed | Attention |
| --- | --- | --- | --- | --- |
| **Hippocampal Volume** | $F(3,310.5)=4.0, p=8.4e-03^*$<br>Non-Hispanic White > Non-Hispanic Black, Latinx [English], Latinx [Spanish] | $F(3,291.0)=3.3, p=0.02^*$<br>Non-Hispanic White > Non-Hispanic Black, Latinx [English] | $F(3,318.1)=5.5, p=1.0e-03^*$<br>Non-Hispanic White > Non-Hispanic Black, Latinx [English], Latinx [Spanish] | $F(3,299.4)=3.5, p=0.02^*$<br>Non-Hispanic White > Non-Hispanic Black, Latinx [English] |
| **Cortical Thickness** | $F(3,280.2)=3.0, p=0.03^*$<br>Non-Hispanic White > Non-Hispanic Black, Latinx [English] | $F(3,263.0)=3.9, p=9.4e-3^*$<br>Non-Hispanic White > Non-Hispanic Black, Latinx [English]<br>Latinx [Spanish] > Non-Hispanic Black, Latinx [English] | $F(3,292.5)=3.8, p=0.01^*$<br>Non-Hispanic White > Non-Hispanic Black, Latinx [Spanish] | $F(3,270.8)=3.5, p=0.02^*$<br>Non-Hispanic White > Non-Hispanic Black, Latinx [English]<br>Latinx [Spanish] > Non-Hispanic Black |
| **White Matter HyperIntensity Volume** | $F(3,283.2)=2.2, p=0.09$ | $F(3,280.9)=2.6, p=0.05^*$<br>Non-Hispanic White > Non-Hispanic Black, Latinx [English] | $F(3,298.8)=2.7, p=0.05^*$<br>Non-Hispanic White > Non-Hispanic Black, Latinx [Spanish] | $F(3,280.9)=2.4, p=0.06$ |
| **Infarcts** | $F(3,345)=4.8, p=2.7e-03^*$<br>Non-Hispanic White > Latinx [English], Latinx [Spanish]<br>Non-Hispanic Black > Latinx [English] | $F(3,169.5)=4.2, p=6.9e-03^*$<br>Non-Hispanic White > Non-Hispanic Black, Latinx [English], Latinx [Spanish]<br>Latinx [Spanish] > Latinx [English] | $F(3,345)=3.9, p=9.4e-03^*$<br>Non-Hispanic White > Latinx [English], Latinx [Spanish]<br>Non-Hispanic Black > Latinx [English], Latinx [Spanish] | $F(3,177.6)=4.6, p=4.1e-03^*$<br>Non-Hispanic White > Non-Hispanic Black, Latinx [English], Latinx [Spanish]<br>Non-Hispanic Black, Latinx [Spanish] > Latinx [English] |

**Supplemental Figure 1.**
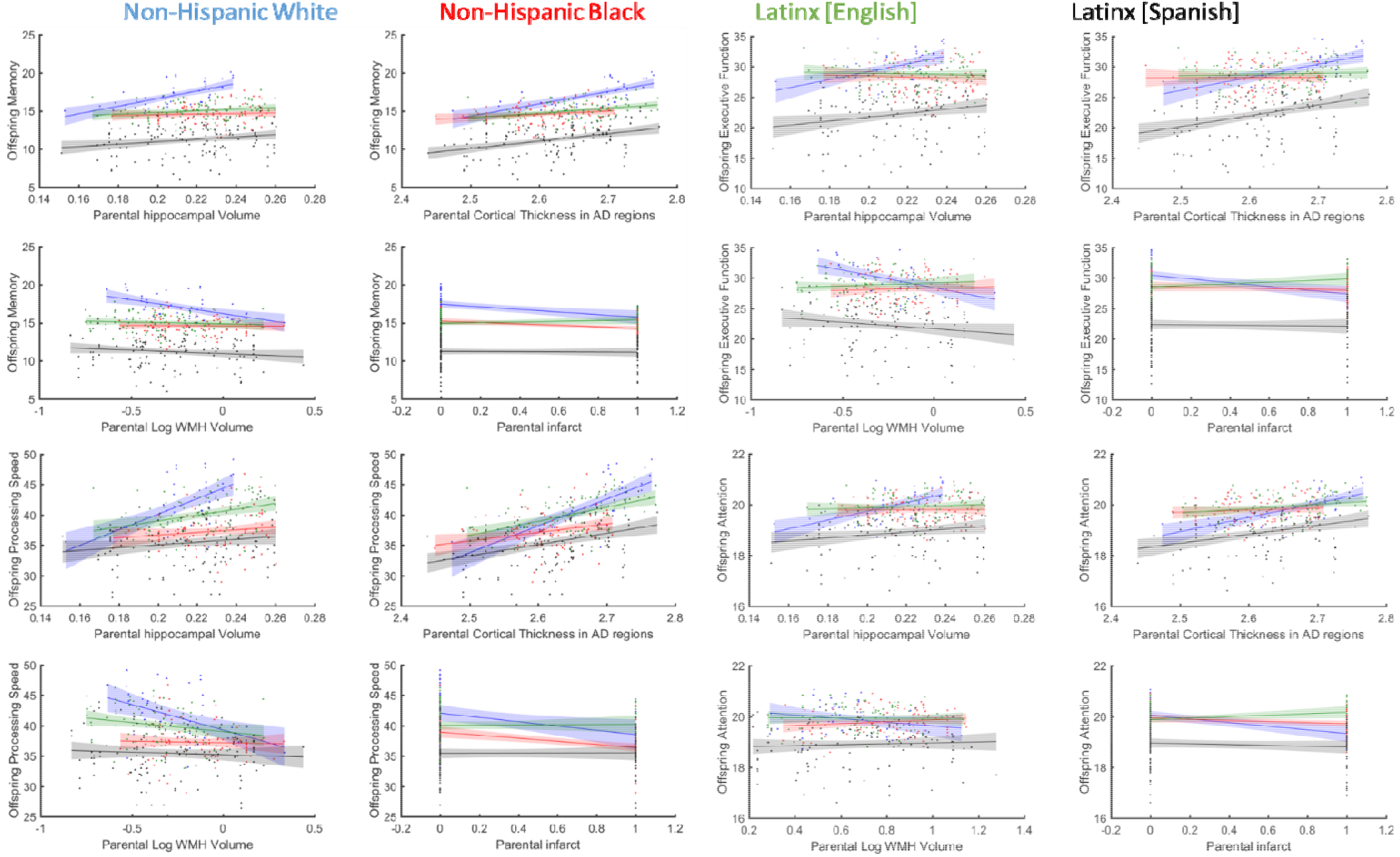
The association of offspring attention scores with parental brain measures by race/ethnicity and language groups.

